# Postoperative throat discomfort and associated factors among elective surgical patients at Ethiopian Teaching Hospitals

**DOI:** 10.1101/2022.09.04.22279584

**Authors:** Hunduma Jisha Chawaka, Zenebe Bekele Teshome

## Abstract

**Introduction:** Throat pain and airway discomfort are the most commonly upsetting conditions of the patients’ wellbeing after surgery. However; little is known about the problem and associated factors.

**Objective:** To assess the magnitude, severity and associated factors of throat discomfort within 24 hours after surgery.

**Design:** **F**acility-based cross-sectional study was employed from July 01 to December 30, 2021. Data was extracted through patient interview and document review during the postoperative period.

**Setting:** The study was conducted at Ambo university referral hospital. On average, about six major elective surgical procedures are performed each day with a total of >1800 patients per year. The patients were scheduled from Orthopedic and trauma, General Surgery, Gynecologic, and Obstetric surgery department.

**Participant:** Using systematic sampling technique 370 elective surgical patients were recruited (k=2) and 36 were excluded from analysis; with incomplete data.

**Result:** The majority 63.2% (211) of the participants were females with a mean age of 35.7 ± 9.7 years. The overall postoperative throat discomfort were observed in 122 (36.5%) and it were severe in 20.5% of them. However; the severity level declined over time 21.9% (21/96), 15.3% (18/118) and 5.6% (5/89); on the 1st, 6th hour and 24th hour, respectively. Utilization of larger ETT (AOR= 6.288, P= 0.001), surgery lasting > 2hours (AOR= 10.839, P= 0.000), awake extubation (AOR= 0.155, P=0.000), frequent episodes of hypotension (AOR= 13.220, P= 0.001), and less experienced anesthetist (AOR= 18.571, P=0.000); were significantly associated with throat discomfort.

**Conclusion:** Post-extubation throat discomfort complaints are high (36.5%). Due attention has to be given to minimize or prevent perioperative frequent episodes of hypotension, higher ETT size utilization, blood loss, and duration of surgery as much as possible. Patient reassurance with analgesia and orientation has to be considered during the postoperative time.

## Introduction

During surgery and anesthesia care, it is supposed to satisfy the demand for patient care and minimize associated complications. A safe and secure airway is achieved by endotracheal tube (ETT), which is performed by an anesthetist or anesthesiologist. In our case, Ethiopia, it is widely done by anesthetists. However, throat pain and airway discomfort is a common, uncomfortable, and distressing sequel after general anesthesia that mainly bores the postoperative patient’s comfort [1, 2]. Many pieces of research have been done worldwide regarding the incidence and factors associated with POST are overwhelmingly varied, ranging from 20 to 70% [3 - 6]. In Ethiopia, few researchers have addressed very few variables and have found the incidence of throat pain is 45.6% in Ethiopia Referral Hospitals [7].

Several studies tried to address a number of possible factors contributing to the development of postoperative throat pain, like being female [8, 9], technical difficulty of intubation, and Suxamethonium use [10], airway suctioning, long duration of surgery, laryngeal masks utilization [11], and or mucosal injury with laryngoscopy, or pharyngeal airway use and larger size endotracheal tubes or cuff pressure [12 - 15].

Many studies revealed being a female has been significantly associated with post-operative throat pain [1, 16 – 18], while others still did not find any difference between the sexes [5, 16, 19].

An existing study often showed highly variable results. As one might expect and know, the current practice of airway management methods and surgical approaches also varies widely from institution to institution. In addition to this, there is limited information about incidence, severity, and contributing factors of postoperative throat discomfort in developing countries like Ethiopia, where there are no adequate resources and qualified manpower.

Furthermore, the stage of extubation estimated blood loss, intraoperative hypotension, cough on ETT, and others have not been considered in the so-far research. In this study, those variables were described once their association with post-operative airway-related discomfort is identified.

Therefore, this study was conducted as a preliminary step to determine the magnitude and severity of postoperative throat discomfort at Ambo university referral hospital and analyze possible factors that contribute to its occurrence.

## Methods and Materials

### Study area setting and Period

This study was conducted at AURH, located 114km away from Addis Ababa to West of Ethiopia, from July 01 to December 30, 2021. The hospital provides a range of health care services through specialty units of surgery, internal medicine, pediatrics, Gynecology and obstetrics, ENT and other minor departments. The hospital supposed to serve for 3.5 million populations and had four major operation theatres (table) on which both emergency and elective surgery is performed. The elective surgery that gets operated on each day ranges from 4 to 10 (on average 6 patients) each day with an approximate more than 1800 surgical procedures in a year. Elective surgeries were scheduled mainly from Orthopedic Surgery, Maxillofacial and dental surgery, Ophthalmology, General Surgery, Gynecologic, and Obstetric specialty units.

All patients were anesthetized using standard intravenous anesthetics (2mg/kg of propofol or 4mg/kg thiopental and 2mg/kg suxamethonium) and maintenance was either halothane or isoflurane. An intubation was made with an endotracheal tube (ETT) size of 6.0 to 7.5 mm internal diameter (ID) tube using standard Macintosh laryngoscope blade.

### Study Design

An institutional-based cross-sectional study was employed.

### Source population

All patients who undergo surgery under general anesthesia during the study period.

### Study population

Patients who undergo elective surgery who met inclusion criteria during the study period.

### Inclusion criteria

All patients whose age is 18 years and or above who undergo surgery will be included in the study.

### Exclusion criteria

Patients who are classified as ASA-3 or above, recent or ongoing upper respiratory tract infection (URTI), patients with a history of recent nasogastric tube insertion or nasal intubation, double-lumen ETT, and pre-existing voice change or difficulty of swallowing were excluded from the study.

### Sample size and Sampling procedure

Using a single population proportion formula and throat pain proportion of 59.6 %; at Gondar University teaching hospital [13]; with 95% CI, 5% margin of error. The sample size calculated to be n = 370

To recruit each study subject we used a systematic sampling technique (k=2). The data were collected from the participant after every two patients. The first patient was arbitrarily selecting from the participants’ list (scheduling board) when the first-day data collection started.

### Study Variable Dependent

- Presence of throat discomfort after elective surgery (yes/no).

### Independent

Sociodemographic characteristics

- Age, sex, BMI Surgical related characteristics
- Surgical diagnosis, duration of surgery/ETT in the trachea, Estimated Blood Loss in milliliter, intraoperative frequency of hypotension episodes and types of surgical procedure,

Anesthetic related characteristics

- ASA classification, duration of anesthesia, ETT size, Extubation Mode (deep/awake), Cough while ETT inside, and Anesthetist’s experience.

### Operational definition

**Throat discomfort** is a sense of any unpleasant feeling or pain at the larynx or pharynx felt during the postoperative time. It was asked with a direct questionnaire survey, ‘Do you have a sore throat after the operation?’ ‘Do you have any pain or difficulty swallowing after the operation?’

**Estimated blood loss** is a total volume of blood lost estimated during surgical procedure which is documented on anesthetic sheet and surgical safety check list.

**Hoarseness** is a harsh or stained voice change that is noticed during a postoperative period by patients. If a nurse observed the patient’s voice change, it was also scored as ‘yes’. A severity level was graded by asking the patient on a visual analogue scale (VAS; 0 no pain, < 3 mild, 4-6 moderate, >7 severe, & 10 the worst pain imaginable).

**Cough** is said to be present (‘YES’) when a sudden, strong abdominal contraction with an attempt/production of cough sound is noticed. It was checked twice, at emergence and during a postoperative time and a single cough was recorded as ‘yes’.

The **presence of hypotension** (HPT) is a drop of blood pressure below 90/60mmHg and recorded once or more times on anesthesia monitoring sheet.

### Endotracheal tube (ETT)

An endotracheal tube is a flexible plastic tube that is placed through the mouth or nose into the trachea to help the patient breathe.

### Laryngoscopy

is a technique of using a device called a laryngoscope to view laryngeal structures to facilitate endotracheal intubation.

### Data Collection

Data was collected through patient interviews and card review using a semi-structured questionnaire during postoperative time; at 1st, 6th hours, and 24 hours, after the patient fully recovered from anesthesia and feels comfortable answering the interviewer’s question. Even though there is no clear time to assess post-operative throat discomfort, it would be better as early as possible, as many scholars suggest [20 – 22]. Demographic data; age, sex, weight, height, American Society of Anesthesiologists (ASA) physical status, surgical procedure type, duration of intubation, size of a cuffed endotracheal tube (ETT) used extubation mode, intraoperative recorded hypotension, estimated intraoperative blood loss (EBL) and anesthetists year of experience were recorded on the prepared sh standard form. The interviewer asks a direct question about dysphagia and or dysphonia-related symptoms, including “choking,” “coughing,” “hard to chew,” “hard to swallow,” “lump in the throat,” and “trouble swallowing,” and any hoarseness or voice change after surgery.

### Data quality control and Analysis Process

A pretest was done on 5% of the sample size before actual data collection and necessary corrections and modifications were made to the questionnaires. Each questionnaire was checked for completeness and entered into SPSS.V.20 statistical software for analysis. Bivariate and multivariate logistic regression analysis was used to determine the association of various factors with the outcome variable. In logistic regression analysis, each explanatory variable with an outcome variable was assessed for its association, and those variables whose p-value < 0.25 were taken as candidate variables for multiple logistic regressions for further analysis of their true association. Variables with a P-value < 0.05 were reported with 95% CI as a statistically significant variable.

### Ethical issue

After an ethical approval were obtained from “Ambo University College of Medicine and Health Science institutional research review board (IRB) committee –with project code of CMHS/S-R-Anesth/02/13”; permission were confirmed from hospital administration. An informed written consent was obtained from each study participant before actual data collection. The confidentiality and privacy of the study participant were kept secret using questionnaire coding. Data was collected after their volunteer participation was confirmed with their signature. Those patients who are diagnosed with having pain were linked to the responsible physician for treatment.

## Result

### Demographic characteristics

Three hundred seventy (370) patients participated from a total of 747 elective surgeries performed during the study period. Of which, 334 (90.3 %) participants’ data were included for analysis (and 36 were excluded because 4 nasal tubes were used, 14 prior throat complaints, and 18 with incomplete documentation). The majority of the participants were female (63.2%), ASA-I (American Society of Anesthesiologists) (77.5%), 18 – 24 BMI (68%), and older than 30 years (66.8%) with a mean of 35.7 ± 9.7 years (**Table 1**).

**Table 1.** Percentage and frequency distribution of Demographic variables of postoperative airway discomfort after extubation, 2021.

| Variables | Classification | Presence of throat discomfort |  | Total |  |
| --- | --- | --- | --- | --- | --- |
|  |  | Yes | No | N | % |
| Sex | Female | 74 | 137 | 211 | 63.2 |
|  | Male | 48 | 75 | 123 | 36.8 |
| Age (year) | Thirty years or less | 43 | 68 | 111 | 33.2 |
|  | More than 30 years | 79 | 144 | 223 | 66.8 |
| BMI | Thirty or more | 6 | 6 | 12 | 3.6 |
|  | 24 – 30 | 28 | 67 | 95 | 28.4 |
|  | 18 – 24 | 88 | 139 | 227 | 68 |
| ASA classification | II | 24 | 51 | 75 | 22.5 |
|  | I | 98 | 161 | 259 | 77.5 |

### Anesthesia and surgery-related characteristics

All patients were anesthetized using standard intravenous anesthetics (2mg/kg of propofol or 4mg/kg thiopental and 2mg/kg suxamethonium) and intubated with an endotracheal tube (ETT) size of 6.0 to 7.5 mm using Macintosh blade laryngoscope. Most surgical procedures lasted more than two hours in 175 (52.4%) and ranged from a minimum duration of 40 minutes to a maximum of 3½ hours. About 46.4% of the participants had recorded intraoperative hypotension episodes; of which 78.7 % (122) had had more than two times episodes. The commonly performed surgery includes general, orthopedics, cesarean delivery, and gynecology in a different position. (**Table 2)**

**Table 2:**
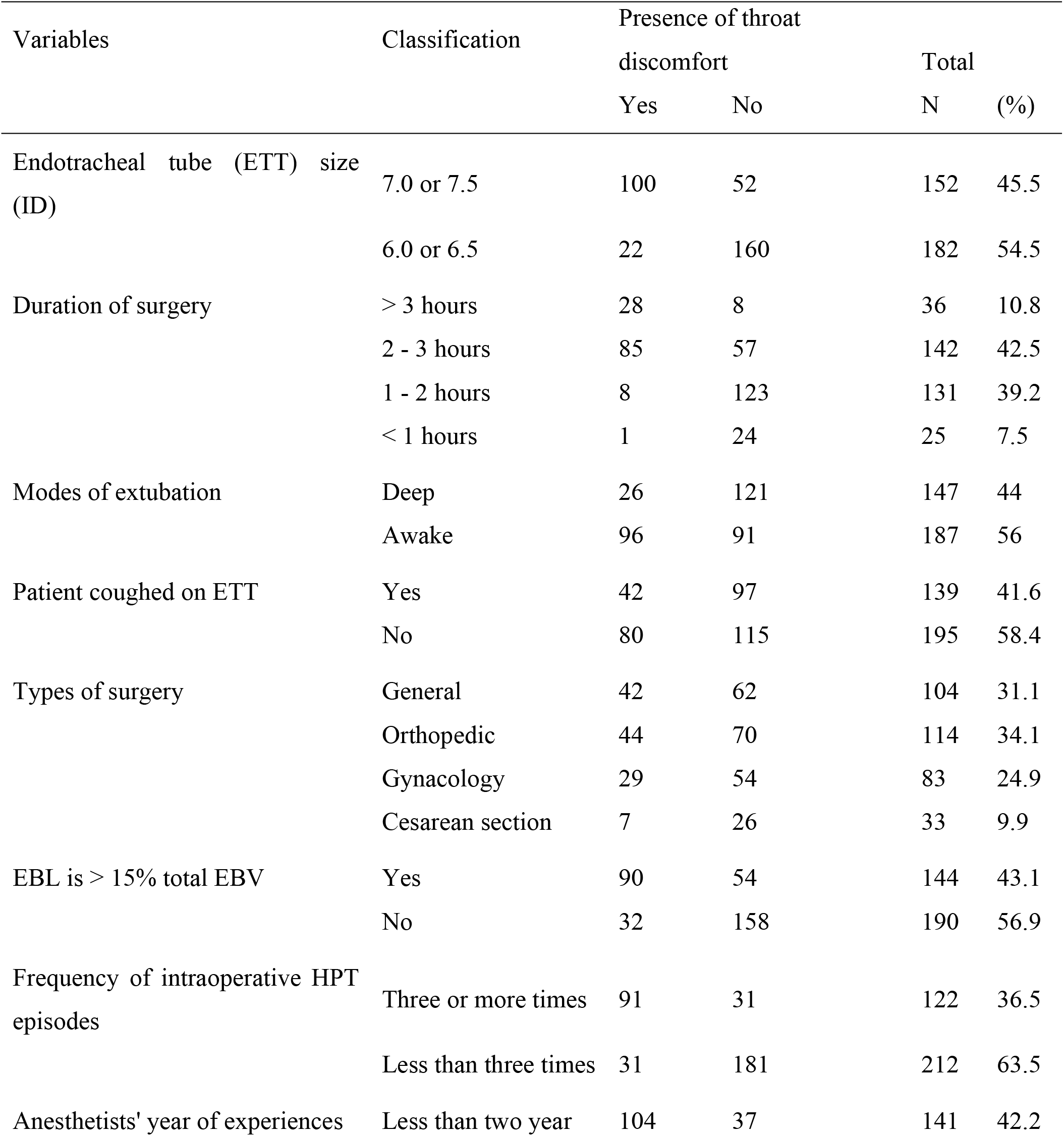
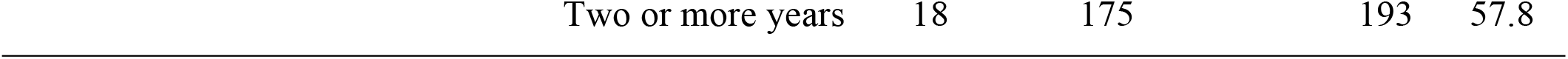
Percentage and frequency distribution of Demographic, Anesthetic and Surgery related variables of postoperative airway discomfort after extubation, 2021.

| Variables | Classification | Presence of throat discomfort |  | Total |  |
| --- | --- | --- | --- | --- | --- |
|  |  | Yes | No | N | (%) |
| Endotracheal tube (ETT) size (ID) | 7.0 or 7.5 | 100 | 52 | 152 | 45.5 |
|  | 6.0 or 6.5 | 22 | 160 | 182 | 54.5 |
| Duration of surgery | > 3 hours | 28 | 8 | 36 | 10.8 |
|  | 2 - 3 hours | 85 | 57 | 142 | 42.5 |
|  | 1 - 2 hours | 8 | 123 | 131 | 39.2 |
|  | < 1 hours | 1 | 24 | 25 | 7.5 |
| Modes of extubation | Deep | 26 | 121 | 147 | 44 |
|  | Awake | 96 | 91 | 187 | 56 |
| Patient coughed on ETT | Yes | 42 | 97 | 139 | 41.6 |
|  | No | 80 | 115 | 195 | 58.4 |
| Types of surgery | General | 42 | 62 | 104 | 31.1 |
|  | Orthopedic | 44 | 70 | 114 | 34.1 |
|  | Gynecology | 29 | 54 | 83 | 24.9 |
|  | Cesarean section | 7 | 26 | 33 | 9.9 |
| EBL is > 15% total EBV | Yes | 90 | 54 | 144 | 43.1 |
|  | No | 32 | 158 | 190 | 56.9 |
| Frequency of intraoperative HPT episodes | Three or more times | 91 | 31 | 122 | 36.5 |
|  | Less than three times | 31 | 181 | 212 | 63.5 |
| Anesthetists' year of experiences | Less than two year | 104 | 37 | 141 | 42.2 |

### Outcome and possible risk factors

Postoperative throat discomfort was complained by 122 (36.5%) participants within 24 hours after extubation. The majority 98(78.8%) of them noticed the discomfort immediately (within an hour); as soon as they got consciousness. (**Figure 1**)

**Figure 1.**
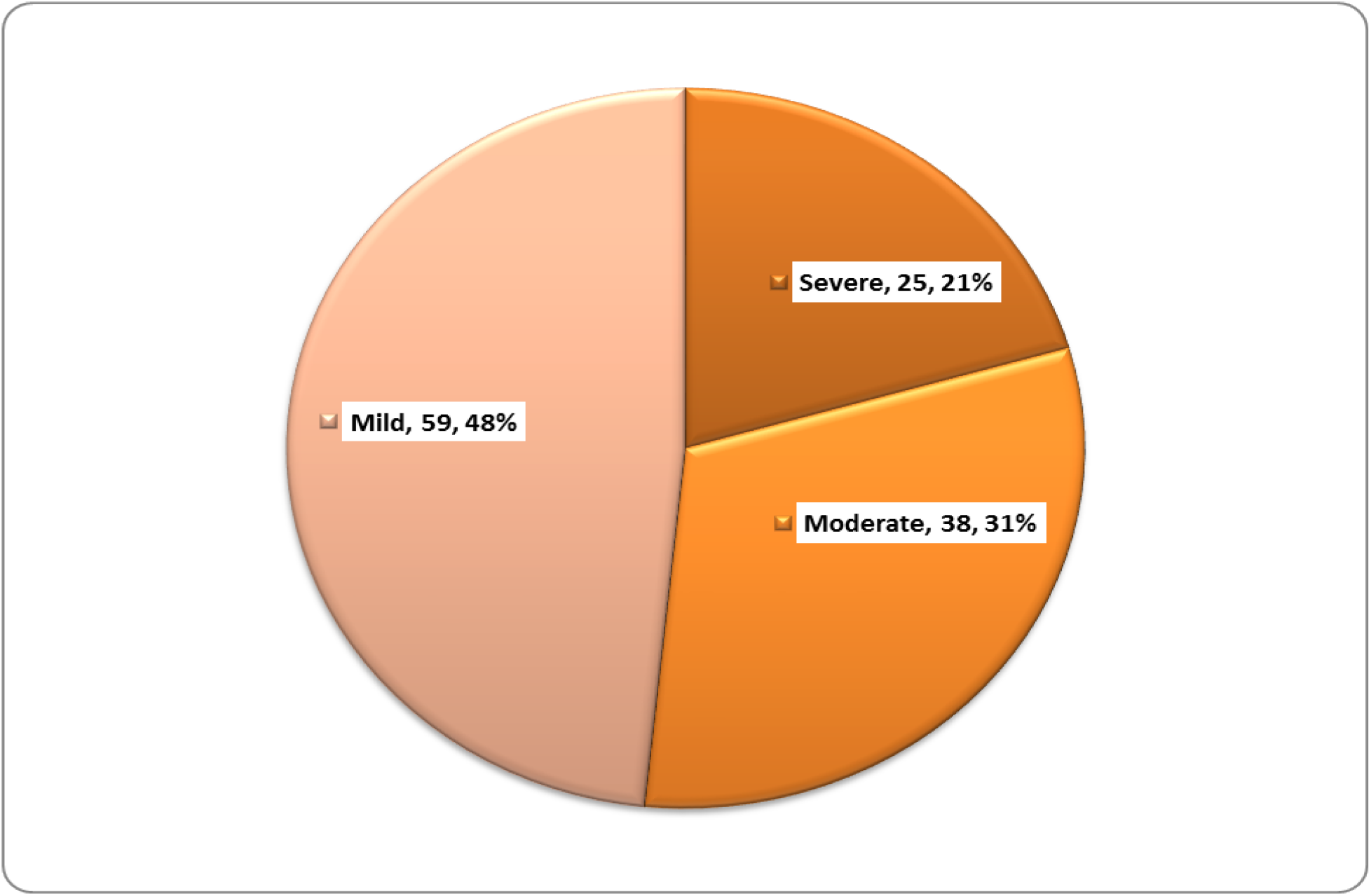
Severity level of postoperative throat discomfort at diagnoses time, 2021.

The overall severity levels were found to be severe in 25(20.5%); moderate in 38(31.1%) and mild in 59(48.4%). However; the severity level of the discomfort seems to decline over time courses. Patients who complain of severe discomfort at 1st (within 1 hour), 2nd (6th hour), and 3rd (24th hour) interviews were found to be 21.9%, 15.3%, and 5.6%; respectively. (**Figure 2**)

The likely hood of throat discomfort complaints among patients extubated in a deep state of anesthesia decreases by 0.16 odd (AOR= 0.16, CI 0.05 – 0.43 P=0.000) compared to awake extubation. Whereas the complaints has positive association with frequent (three or more) episodes of intraoperative hypotension (AOR= 12.85, CI 4.56 – 36.18, P= 0.000), after a larger size of an endotracheal tube (ETT) (AOR= 6.26, CI 2.13 – 18.43, P= 0.001), the surgery lasted two or more hours duration (AOR= 11.20, CI 3.53 – 35.50, P= 0.000), and less experienced (< 2years) anesthetists by 18.10 more when compared to the counterpart patients (AOR= 18.10, CI 6.51 – 50.31, P=0.000). (**Table 3**)

**Table 3:** Logistic regression output of associated variables of postoperative airway discomfort after extubation, 2021.

| Variables | COR | AOR | CI (95%) | P-value |
| --- | --- | --- | --- | --- |
| Frequent episode of intraop hypotension | 3.60 | 12.85 | 4.56 – 36.18 | .000 |
| Larger endotracheal tube size (7.0 -7.5) | 6.46 | 6.26 | 2.13 – 18.43 | .001 |
| Duration of surgery lasting 2 or more hours | 12.47 | 11.20 | 3.53 – 35.50 | .000 |
| Deep extubation | 0.15 | 0.16 | 0.05 – 0.43 | .000 |
| Estimated blood loss > 15% of estimated blood volume | 2.83 | 2.46 | 0.92 – 6.62 | .084 |
| Anesthetists' experience less than two years | 20.13 | 18.10 | 6.51 – 50.31 | .000 |

## Discussion

Achieving a safe and secure airway by endotracheal tube (ETT) plays a vital role during anesthesia care services. However; airway-related complaints after surgery under general anesthesia has been continued to be challenging and overwhelming the comfort patients during postoperative time Sore throat, hoarseness, dysphagia, neck pain, difficulty to cough, and dysphonia are among the commonest problems after endotracheal tube extubation (ETT) [2, 3, 23].

Anesthesia science and practices had advanced too much distance in decreasing postoperative morbidity and mortality; within the last few decades. However; the problem is much worse in sub-Saharan countries.

The prevalence of postoperative airway-related complaints is widely varied, ranging from 20 to 70% [3 – 6]. In developed countries universities or teaching hospitals in Iran [24], Saudi Arabia [25] and China [26]; the incidence is (documented as low as 26.3%, 36.3% and 16.2 – 31.9%, respectively. Whereas In Nigeria teaching hospital it was found to be as high as 63% [5] and 58.9% [27]; In Ethiopia, the two studies conducted in the northern part of the country at Gondar and Debretabor teaching hospital showed, 59.6% and 48.8%; respectively [13, 28].

Here in Ambo University referral hospital, we found the overall prevalence of post-operative throat discomfort after 24 hours is 36.5%. Our study relatively found less prevalence than the others. However, incongruent with our result a study conducted in a Turkey teaching hospital found 37.7% throat pain complaints [2]. This variation could be explained as the inconsistent standard of care followed during perioperative time; which is challenging in resource-limited settings like AURH, where the burden of illness and mortality from potentially treatable conditions is higher than in resource-rich settings where standards of operating procedures guidelines are implemented. Barriers to providing quality care in different settings include a lack of epidemiological data and contextual evidence for medical decision-making, gaps in health systems organization and resources, and institutional barriers to implementation. On top of this, the discrepancy might be due to sample sizes and data collection time differences. On the other hand, the educational level and understanding level of the question to be replied might be responded to differently.

In this study, we found larger endotracheal tube (7.0 -7.5) is associated (AOR=6.26, 95% CI=2.13-18.43, p=0.001) with postoperative throat discomfort. A different researcher had reported similar findings [16, 18, 24, 26].

This is explained for two reasons; direct mechanical trauma and prolonged vascular compression resulting in low blood flow to the mucosa that might be followed by the inflammation process. With similar justification, prolonged surgery (two or more hours) duration of procedures (AOR=11.20, 95% CI=3.53 – 35.50, p=0.00), and less experienced (less than 2years) anesthesia provider (AOR=18.10, 95% CI=6.51 – 50.31, p=0.00) were found to be associated with the postoperative airway related discomfort and complain. This is because more manipulation and repeated attempts at laryngoscopy are common in less experienced anesthetists. On the other hand, long-standing compression of the underlying laryngeal mucosal surfaces by inflated EET cuff pressure is accompanied by low perfusion to the area and mucosal injury from hypo perfusion. Incongruent with our findings, the different researchers explained this scenario [5, 6, 17]. Unlike other studies, we did not find an association between gender and throat discomfort. Many other researchers also found no difference in sore throat complaints between males and females [24]. In the future, this area needs to be studied in-depth.

This study depicts that the severity of complaint of throat discomfort decreases over time; on the 1st Vs 6th, Vs 24th-hour interviews 21.9%, 15.3%, and 5.6%, respectively complained of a severe form of discomfort. This finding is supported by Farazmehr et al.; they found that the prevalence of sore throat post laryngeal mask use within 1, 6, and 12 h after the surgery was 16.5 %–36.1 %, 13.6 %– 33.3 %, and 10.7 %–28.7 %, respectively [24]. There are also other researchers who identified that throat pain is declining after 6 hours [25, 29].

We observed that some participants were recovered within 6hours (two individuals) and 33 patients within 24 hours. This can be explained by the swelling secondary to inflammation becoming resolved as time elapsed among patients whose inflammation is not severe enough [30].

### The implication of the paper

As one might expect and know, current practice of airway management methods and surgical approaches widely varies from institution to institution. This paper will widen the insight of emphasis to be given to the preventable risk factors of postoperative throat discomfort specially in the limited resource countries. It will help to prevent or minimize postoperative associated complications and mortality.

### Limitation of the study

Although this study provided valuable information, there are some limitations that could be addressed in another study. A chart review might be retrieved with incomplete information and the patients were interviewed at an inconvenient time. In addition, data on other variables needed to observe intactness of recurrent laryngeal nerve injury, vocal cord examination and ETT cuff pressure monitoring during the intraoperative period; to predict whether the throat discomfort resulted from airway instrumentation or direct surgical insults during procedure; were not addressed due to the impossibility of the resource available and accesses in our setup.

### Conclusion and recommendation

During anesthesia care, securing an airway using an endotracheal tube is a very important part of care. However, throat pain and airway discomfort continue to be upsetting conditions of patient comfort after surgery in resource-limited countries. The overall postoperative throat discomfort complaint within 24 hours after endotracheal tube (ETT) extubation was high (36.5%) and it was severe at 21.3% (26/122)

Due attention has to be given in the case of larger ETT-sized utilization, surgery lasting > 2hours duration, awake extubation, frequent episodes of intraoperative hypotension, and less experienced anesthetist.

## Data Availability

All relevant data are within the manuscript and can be available on request

## Conflict of interest

We, authors, declare there is no conflict of interest; authorship, grant, and or for result.

## Source of funding

no

## Authors’ contribution

All authors (HJ, and ZB) have worked from the initiation of study design and the interpretation of data. Both authors participated in the writing, critically revised and approve the final manuscript.

## Acknowledgement

We would like to thank all participants who gave us their precious time during data collection. Our appreciation extended to Ambo University referral hospital officials and staff for their cooperation and facilitation. Our gratitude also extends to the anesthesia team for their tireless assistance in making this research possible by delivering valuable information in all aspects.

## Notes

### Competing Interest Statement

The authors have declared no competing interest.

### Funding Statement

The author(s) received no specific funding for this work.

### Author Declarations

Approved by Ambo University College of Medicine and Health Science institutional research review board (IRB) committee –with project code of CMHS/S-R-Anesth/02/13”

